# Correlates of protection, viral load trajectories and symptoms in BA.1, BA.1.1 and BA.2 breakthrough infections in triple vaccinated healthcare workers

**DOI:** 10.1101/2022.04.02.22273333

**Authors:** Ulrika Marking, Sebastian Havervall, Nina Greilert Norin, Oscar Bladh, Wanda Christ, Max Gordon, Henry Ng, Kim Blom, Mia Phillipson, Sara Mangsbo, Anna Smed Sörensen, Peter Nilsson, Sophia Hober, Mikael Åberg, Jonas Klingström, Charlotte Thålin

**Affiliations:** Department of Clinical Sciences, Karolinska Institutet Danderyd Hospital, Stockholm, Sweden; Center for Infectious Medicine, Department of Medicine Huddinge, Karolinska Institutet, Stockholm, Sweden; Department of Medical Cell Biology and SciLifeLab, Uppsala University, Uppsala, Sweden; Department of Pharmacy and SciLifeLab, Uppsala University, Uppsala, Sweden; Division of Immunology and Allergy, Department of Medicine Solna, Karolinska Institutet, Karolinska University Hospital, Stockholm, Sweden; Department of Protein Science, KTH Royal Institute of Technology, SciLifeLab, Stockholm, Sweden; Department of Medical Sciences, Clinical Chemistry and SciLifeLab, Uppsala University, Uppsala, Sweden

## Abstract

**Background:** Booster vaccine doses offer protection against severe COVID-19 caused by omicron but are less effective against infection. Characteristics such as serological correlates of protection, viral abundance, and clearance of omicron infection in triple vaccinated individuals are scarce.

**Methods:** We conducted a 4-week twice-weekly SARS-CoV-2 qPCR screening shortly after an mRNA vaccine booster in 368 healthcare workers. Spike-specific IgG levels and neutralization titers were determined at study start. qPCR-positive participants were sampled repeatedly for two weeks and monitored for symptoms.

**Result:** In total 81 (cumulative incidence 22%) omicron infections were detected, divided between BA.1, BA.1.1 and BA.2. Increasing post-booster antibody titers were protective against infection (p<0.05), linked to reduced viral load (p<0.01) and time to viral clearance (p<0.05). Only 10% of infected participants remained asymptomatic through the course of their infection. Viral load peaked at day 3 and live virus could be detected for up to 9 days after first PCR-positive sample. Presence of symptoms correlated to elevated viral load (p<0.0001), but despite resolution of symptoms most participants showed Ct levels <30 at day 9. No significant differences were observed for viral load and time to viral clearance between BA.1, BA.1.1 and BA.2 infected individuals.

**Conclusion:** We report a high incidence of omicron infection despite recent booster vaccination in triple vaccinated individuals. Increasing levels of vaccine-induced spike-specific WT antibodies entail increased protection against infection and reduce viral load if infected. High viral load and secretion of live virus for up to nine days may facilitate transmission in a triple vaccinated population.

## Introduction

The SARS-CoV-2 B.1.1.529 (omicron) variant has caused a considerable surge in COVID-19 cases, including in populations with high vaccine uptake [1]. While the now widely administered booster mRNA vaccine (third dose) have been shown to be effective against severe COVID-19 [1 2] caused by omicron, protection against infection appears limited and not sufficient to prevent viral transmission [3 4]. Vaccine induced serological responses correlated well with the risk of infection with the ancestral virus and pre-omicron SARS-CoV-2 variants of concern [5-8], but less is known regarding the correlation between serological response and protection against omicron infection.

The omicron surge was initially caused by sublineages including BA.1, BA.1.1 and BA.2 [9]. BA.2 carried a transmission advantage, taking over as the dominating sublineage in several countries [10] and has gradually been replaced by other substrains [11]. Mutations in the antibody target spike protein distinguish the omicron sublineages from each other, but *in vitro* neutralization data suggest similar vaccine induced neutralizing capacity against BA.1 and BA.2 [12]. Viral characteristics and variations in viral abundance and clearance between the sublineages are, however, not fully characterized [13-15].

Here we investigated breakthrough infections in triple-vaccinated healthcare workers (HCW) with and without prior SARS-CoV-2 infection. During the study period BA.1, BA.1.1 and BA.2 circulated in Stockholm, Sweden, allowing for comparison of breakthrough infections with the three sublineages [9]. Serological correlates of protection against infection, symptoms and viral RNA trajectories were analyzed.

Cumulative incidence over the study period was 22%, despite a recent booster vaccine dose, and viral RNA trajectories were similar in BA.1, BA.1.1 and BA.2 and suggestive of infectivity. Although increasing post booster antibody titers entailed a protective role against infection and had a reducing effect on viral load, these data imply a substantial evasion of vaccine induced immunity.

## Methods

### Study cohorts

The COMMUNITY study comprises 2149 HCW at Danderyd Hospital, Stockholm, Sweden, enrolled between April and May, 2020. Study participants are followed every four months since inclusion [16-19] with blood samples and collection of relevant information (such as chronic medication/immune suppression, work related SARS-CoV-2 exposure, etc.) through a smart phone-based application. SARS-CoV-2 infection prior to vaccination was confirmed by seroconversion at any of the follow-up visits and/or by PCR. All HCW were offered vaccination with either BNT162b2 (BNT) or ChAdOx1 nCoV-19 (ChAd), depending on availability, starting in January 2021. Data regarding which vaccine and date of vaccination is obtained through the Swedish vaccination register (VAL Vaccinera) and data regarding PCR-confirmed SARS-CoV-2 infection is obtained through the national communicable disease surveillance register SmiNet (Swedish Public Health Agency). The current sub-study utilizes samples taken from two overlapping cohorts, both derived from the COMMUNITY study cohort (Figure 1A).

**Figure 1.**
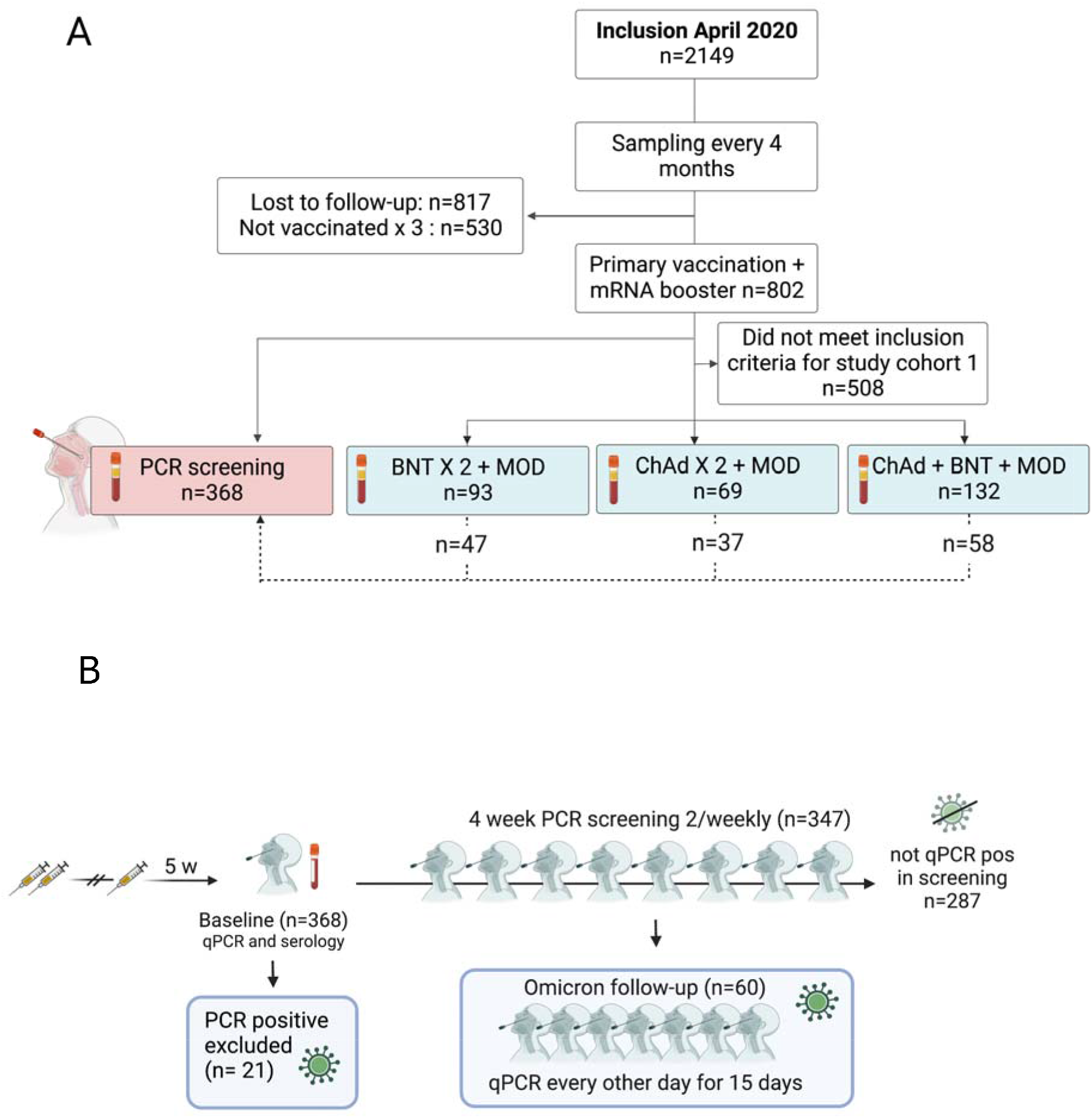
(**A**) Overview of COMMUNITY study cohort and cohorts in this sub study. For investigation of serological responses to third vaccine dose *study cohort 1* (blue) was used. For qPCR screening of recently boosted individuals, *study cohort 2* (red) was used. (**B**) Overview of PCR screening study. Nasal/oropharyngeal/saliva swabs for PCR and serum were collected at enrolment (baseline). Participants who tested qPCR positive at enrolment (n=21) were excluded from the screening. Self-administered nasal/oropharyngeal/saliva swabs for qPCR were collected twice weekly for 4 weeks or until positive qPCR test from participants who tested qPCR negative at enrolment (n=347). Participants who subsequently tested qPCR positive during the screening program (n=60) were enrolled in an extended program comprising self-administered nasal/oropharyngeal/saliva swabs for qPCR every other day for 15 days.

To investigate serological responses to an mRNA-1273 (MOD) booster dose, we analyzed samples taken at the sixth follow up on January 13^th^ - Feb 7^th^ 2022 (*cohort 1, figure 1A*). Participants with a PCR-confirmed SARS-CoV-2 infection between first vaccination and up to 6 days after post booster blood sampling (n=40) were excluded from *cohort 1* to limit interference of breakthrough infections. In order to further limit interference of variations in dose intervals and time between vaccination and sampling, we selected samples from all participants who had received a MOD booster dose between December 13 – 23 2021, after either 1. primary vaccination with two doses of BNT (dose interval 33-53 days with second dose between May 23 and June 30, 2021, n=93), 2. two doses of ChAd (dose interval 70-108 days with second dose between May 10 and June 14, 2021, n=69) or 3. one dose of ChAd followed by one dose of BNT (dose interval 68-115 days with second dose between May 18 and June 21, 2021, n=132). Participants were stratified by occurrence of SARS-CoV-2 infection prior to primary vaccination. The analyzed samples were collected 13 weeks (median 90 days, IQR 82-98 days) after second vaccine dose and 5 weeks (median 35 days, IQR 33-40 days) after the booster dose. Demographics for this cohort (*cohort 1*, Figure 1A) are presented in Table 1.

**Table 1.** Demographics, frequency of prior SARS-CoV-2 infection and vaccine regimen of *study cohort 1*: the investigation of serological booster responses. IQR; interquartile range; ChAd; ChAdox1 n-CoV 19 vaccine, BNT; BNT162b2 mRNA vaccine, MOD; mRNA-1273 vaccine.

|  | BNTx2 + MOD<br>(N=93) | ChAd+BNT + MOD<br>(N=132) | ChAdx2 + MOD<br>(N=69) |
| --- | --- | --- | --- |
| **Age** |  |  |  |
| Median (IQR) | 51 (40-59) | 51 (45-57) | 52 (47.5-58.5) |
| **Sex** |  |  |  |
| Female | 84 (90%) | 115 (87%) | 61 (88%) |
| Male | 9 (10%) | 17 (13%) | 8 (12%) |
| **Prior SARS-CoV-2 infection** |  |  |  |
| No | 18 (19%) | 86 (65%) | 34 (50%) |
| Yes | 75 (81%) | 46 (35%) | 35 (50%) |

To investigate risk of and viral characteristics of omicron breakthrough infections in triple vaccinated participants, we invited 368 participants to a twice-weekly qPCR screening with self-administered naso-oropharyngeal/saliva swabs [20] for four weeks. Inclusion to this screening study was conducted in conjunction to the sixth follow-up in January 2022. All participants who had completed primary vaccination and received a BNT or a MOD booster were invited (n=802), of which the first 368 participants who came to the sixth follow-up and conceded to the screening study were included. This cohort (*cohort 2*) therefore overlaps with the cohort chosen to investigate serological responses to an mRNA-1273 (MOD) booster dose (*cohort 1*), Figure 1A. Participants with qPCR confirmed SARS-CoV-2 infection between their third vaccine dose and appointment for study inclusion were not included in the PCR screening study (*cohort 2*). Participants testing positive at the first qPCR test in the screening study were excluded from further analysis but included in estimation of cumulative incidence. Demographics for this cohort (*cohort 2*) are presented in Table 2. Positive qPCR tests after a negative inclusion test were followed by an extended set of self-administered swabs for qPCR every second day for 15 days post first positive sample. All study participants who engaged in the extended follow-up responded to a questionnaire including a pre-defined set of symptoms (fever, sore throat, cough, headache, anosmia and rhinorrea). After completing 15 days of follow-up sampling, participants continued in the twice-weekly screening until the end of the study period. PCR, whole genome sequencing and virus isolation was performed as previously described [21]. The screening study procedure is presented in Figure 1B.

**Table 2.** Demographics, frequency of prior SARS-CoV-2 infection and booster vaccine regimen of *study cohort 2*: the qPCR screening. Pos; positive, IQR; interquartile range, MOD; mRNA-1273 vaccine, BNT; BNT162b2 mRNA vaccine

|  | Omicron pos<br>(N=60) | Not Omicron pos<br>(N=287) | All<br>(N=347) |
| --- | --- | --- | --- |
| **Age** |  |  |  |
| Median (IQR) | 51 (40-58) | 54 (46-60) | 53 (45-60) |
| **Sex** |  |  |  |
| Female | 55 (92%) | 255 (89%) | 310 (89%) |
| Male | 5 (8%) | 32 (11%) | 37 (11%) |
| **Prior SARS-CoV-2 infection** |  |  |  |
| No | 43 (72%) | 160 (56%) | 203 (58%) |
| Yes | 17 (28%) | 127 (44%) | 144 (42%) |
| **Booster vaccine** |  |  |  |
| MOD | 45 (75%) | 218 (76%) | 263 (76%) |
| BNT | 15 (25%) | 69 (24%) | 84 (24%) |
| **Days 3rd vaccine to inclusion** |  |  |  |
| Median (IQR) | 34 (31-36) | 34 (32-37) | 34 (32-37) |

The study was approved by the Swedish Ethical Review Authority (dnr 2020-01653) and conducted in accordance with the declaration of Helsinki. Written informed consent was obtained from all study participants.

### Serological investigation

SARS-CoV-2 WT spike-specific IgG and cross-reactive IgG capable of binding omicron (BA.1, BA.1.1 and BA.2) were measured in post vaccination samples drawn at start of the screening study (V-PLEX SARS-CoV-2 Panel 22, 23 and 25, Meso Scale Diagnostics, Maryland, USA). Antibody titers are expressed as arbitrary units (AU)/ml, except for WT spike-specific IgG, which are expressed in the WHO-standard binding antibody units (BAU)/ml.

### Live-virus microneutralization

In order to establish correlations to binding antibody assays, a sub-set of participants were tested for neutralizing capacity against SARS-CoV-2 WT and omicron BA.1 using a micro-neutralization assay as previously described [22]. All samples included in both cohorts were stratified by primary vaccine regimen and 64 (maximum capacity) samples randomly selected, (45 SARS-CoV-2 naïve and 19 recovered). Briefly, heat-inactivated serum was 3-fold serially diluted, mixed with virus, incubated for 1 hour and finally added, in duplicates, to confluent Vero E6 cells in 96-well plates. Original SARS-CoV-2 WT and omicron BA.1 (both isolated from Swedish patients) were used. After 5 days incubation, the wells were inspected for signs of cytopathic effect (CPE) by optical microscopy. Serum neutralizing activity was measured by 100% CPE inhibition (IC100). Each well was scored as either neutralizing (if no signs of CPE was observed) or non-neutralizing (if any CPE was observed). The arithmetic mean neutralization titer of the reciprocals of the highest neutralizing dilutions from the two duplicates for each sample was then calculated.

### Virus isolation

All respiratory samples were added to confluent Vero E6 cells and incubated for 10 days with regular exchange of medium. Viral infectivity was assessed manually by microscopy (signs of CPE) and was confirmed by qPCR (lower Ct-value in supernatants over time, showing active viral replication).

### Statistics

Risk of breakthrough infection over the four weeks screening period was evaluated using a Poisson regression model with a log offset for observed time. Time was defined as weeks until infection or study end. We investigated the change in risk of infection for every log2 increase (i.e. two-fold increase) in antibody level, while adjusting for prior infection status, age and sex. There was no support for the need of applied interaction between prior infection status and antibody levels (p>0.9). Linear regression models were generated for nadir Ct levels and for number of days until negative qPCR, both adjusted for age, sex, prior infection status, log2-transformed WT spike-specific serum IgG levels and occurrence of asymptomatic course of infection. Since whole genome sequencing was performed only if Ct <30, Ct variations between different strains were investigated in separate regression models. For statistical comparisons, negative qPCR-samples were given a Ct value of 46. Mann-Whitney U test was performed for comparisons of antibody titers between groups, correlations were estimated by non-parametric Spearman correlation test. Statistical analyses were performed using GraphPad Prism version 9.2.0 (GraphPad Software, San Diego, California, USA) or statistical program R version 4.2.1.

## Results

### Serological responses following MOD booster vaccine dose

We have previously reported increased cross-neutralization potency against SARS-CoV-2 variants following primary vaccination with two doses BNT or heterologous ChAd followed by BNT as compared to two doses of ChAd [23]. Using blood samples taken before (figure 2A) and five weeks after (figure 2B) MOD booster vaccination, we found comparable post booster anti-WT spike IgG (figure 2B), anti-WT RBD IgG (figure S1A), cross-reactive IgG capable of binding omicron BA.1 spike (figure S1B) and RBD (figure S1C) and neutralizing titers against both WT (figure S1D) and omicron BA.1 (figure S1E). As previously reported

**Figure 2.**
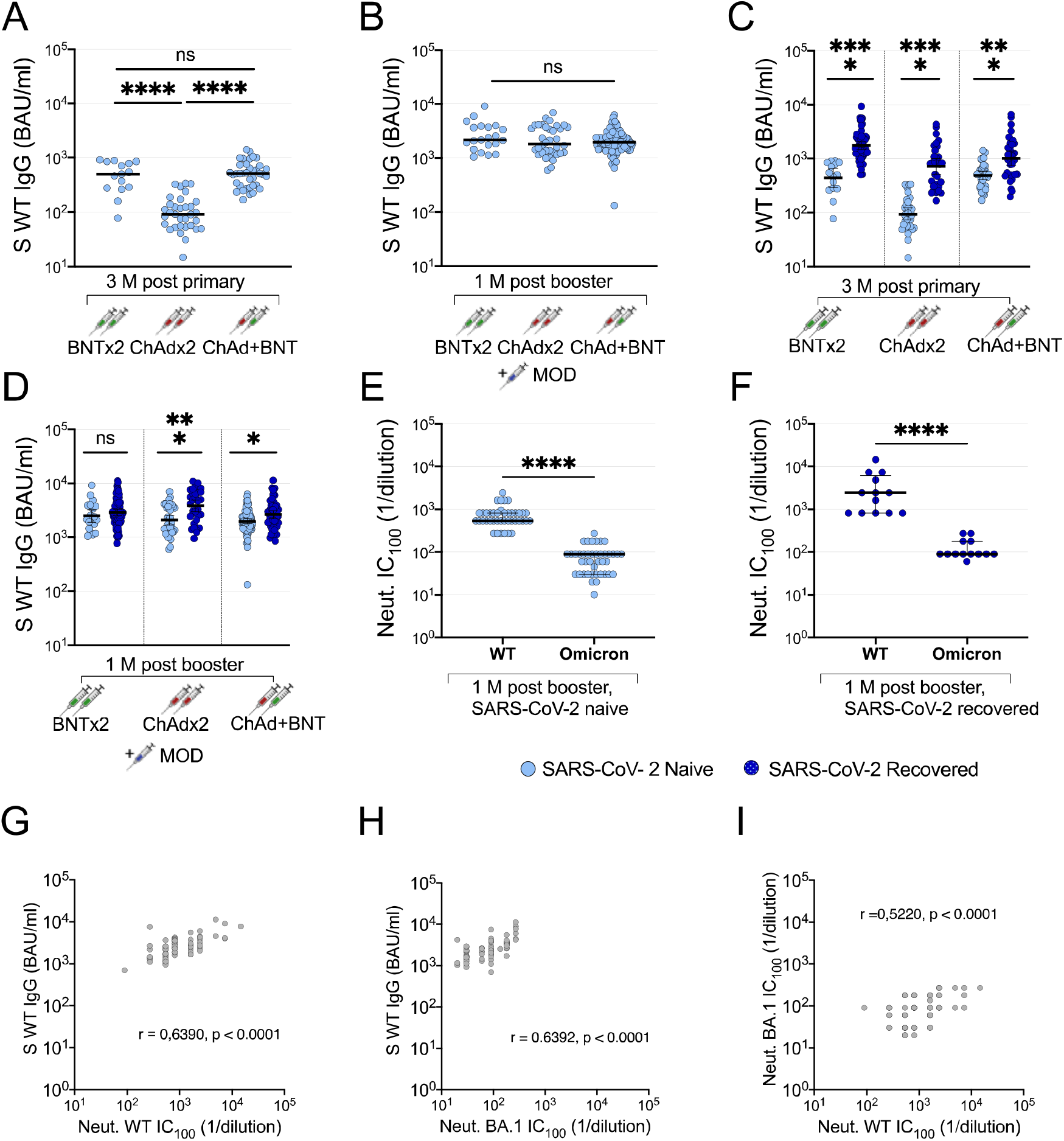
Impact of primary vaccine regimens and prior infection on booster antibody responses. Anti-WT spike IgG three months after primary vaccination series with two doses BNT, two doses ChAd or one dose ChAd followed by one dose BNT (**A)**, and anti-spike IgG one month after MOD booster vaccine dose in the same participants (**B)**. Anti-WT spike IgG in SARS-CoV-2 naïve (light blue dots) and recovered (dark blue dots) participants 3 months after primary vaccination series with two doses BNT, two doses ChAd or one dose ChAd followed by one dose BNT (**C)** and one month after MOD booster vaccine dose in the same participants (**D)**. Microneutralizing titers against SARS-CoV-2 WT and omicron BA.1 in SARS-CoV-2 naïve (**E**) and recovered (**F**) participants one month after MOD booster vaccine dose. Correlation of spike-specific IgG titers to live virus neutralization with SARS-CoV-2 WT (**G**) and omicron (**H**), and correlation of SARS-CoV-2 WT and omicron strains in live virus neutralization (**I**). IgG titers are presented as binding antibody units (BAU)/ml and microneutralizing titers are presented as lowest neutralizing dilution (1/Y). Lines depict geometric mean titers and bars depict 95% confidence interval. S; spike, WT; wild-type, Neut; microneutralizing titer, BAU; binding antibody units; BNT; BNT162b2 mRNA vaccine, ChAd; ChAdox1 n-CoV 19 vaccine, MOD; mRNA-1273 vaccine, M; months, ns; P > 0.05, *; P ≤ 0.05, **;P≤0.01,***;P≤0.001, ****; P ≤ 0.0001

[24] SARS-CoV-2 recovered vaccinees showed stronger antibody responses than SARS-CoV-2 naïve vaccinees following primary vaccination (Figure 2C). This difference still remained after the booster dose (Figure 2D), although not as strong (fold change 2.5 vs. 8.2 for anti-spike WT IgG (Figure 2D), 1.3 vs. 4.5 for cross-reactive IgG capable of binding Omicron BA.1 spike (Figure S1F), 2.7 vs. 7.6 for neutralizing titers against WT (Figure S1G) and 1.5 vs. 18 for neutralizing titers against Omicron BA.1 (Figure S1H) after booster dose as compared to after primary vaccination).

Consistent with recent reports [25-27], post booster neutralization of omicron BA.1 was substantially lower compared to neutralization of WT both in SARS-CoV-2 naïve participants (10.2 fold-change, p<0.001, Mann-Whitney U-test) (Figure 2E) and in SARS-CoV-2 recovered participants (fold change 19.2, p<0.0001, Mann-Whitney U-test) (Figure 2F). Correlation between WT spike-specific IgG and live virus microneutralization of ancestral and omicron SARS-CoV-2 were equally high, both spearman r 0.64, p<0.0001 (Figure 2G-H). Although omicron neutralization titers were significantly lower, live WT SARS-CoV virus neutralization correlated to vs BA.1 sublineage neutralization (spearman r 0.52, p<0.0001) (Figure 2I).

Taken together, these findings show that while booster vaccination may level out differences due to various prior immunizations and infection history, the serological response against omicron is clearly lower than against WT SARS-CoV-2.

### High rate of Omicron breakthrough infection in triple-vaccinated HCW January – February 2022

To assess the risk of breakthrough infections with omicron following booster vaccination, 368 triple-vaccinated participants were enrolled in a qPCR-screening study early 2022. Self-administered naso-oropharyngeal/saliva tests were performed twice weekly for four weeks (median adherence 2 samples per week, IQR 1.75-2). The total time-at-risk was 1324 person-weeks, total number of screening samples 2068.

A total of 81 (22% of all included participants (n=368)) omicron breakthrough infections were detected during the four-week screening period, among which 21 were detected at study inclusion and excluded from further analysis but the estimation of cumulative incidence. Frequency of work with COVID-19 patients, work with non-COVID-patients or non-patient related work was similar among participants with and without omicron breakthrough infection (Table S1). 60 participants who were negative at first sample and subsequently tested positive during the four-week screening were enrolled in a 14-day follow up with self-administered samples every second day. Adherence to follow-up samplings was high with a median of 7 (IQR 7-7) self-administered follow-up samples. Viral RNA levels reached peak levels at day three after the initial positive test, and the majority of the participants remained positive with Ct < 30 nine days after initial positive test (Figure 3A).

**Figure 3.**
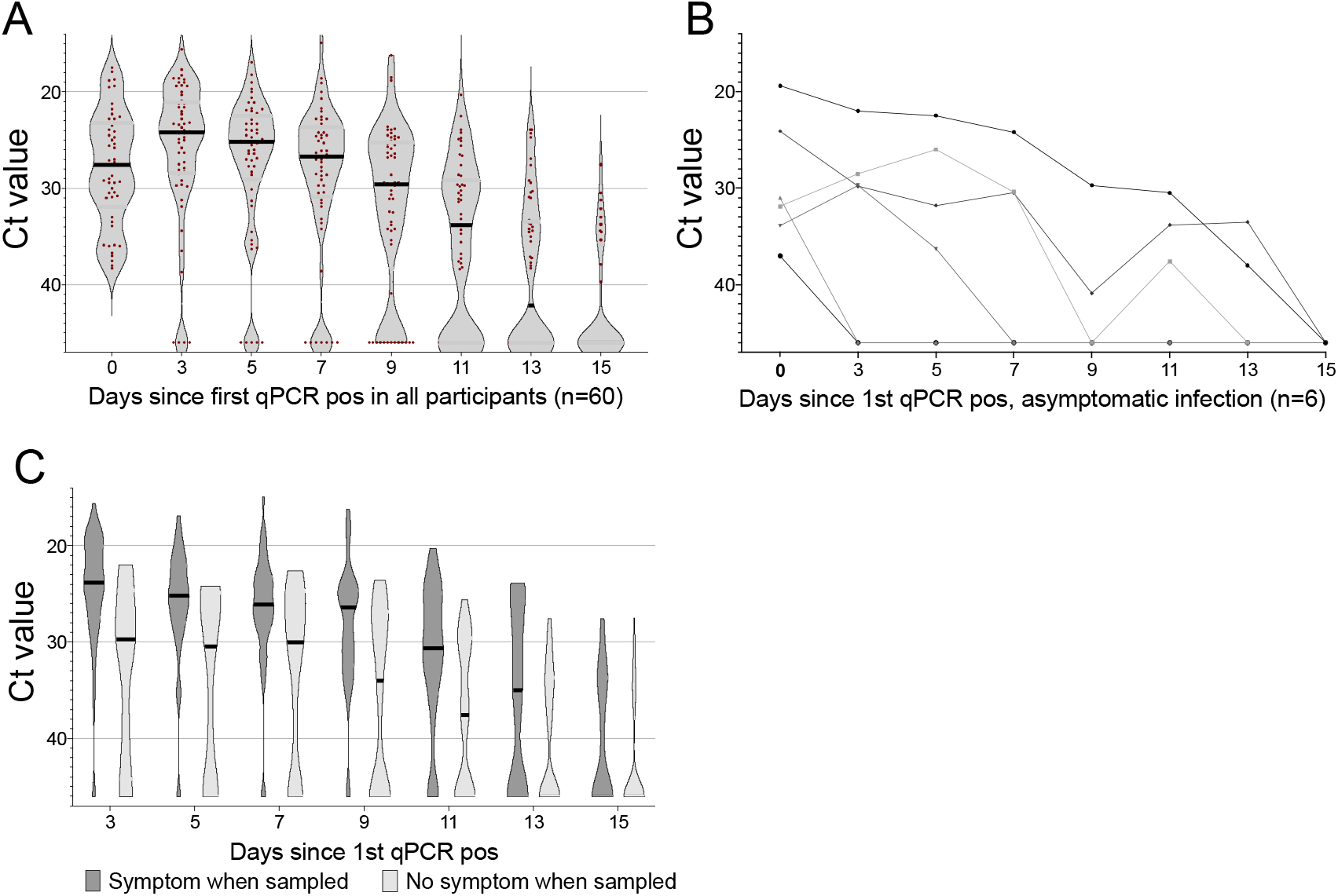
SARS-CoV-2 omicron viral load over the first 15 days of infection, Ct relation to symptoms and pre-infection antibody titers in participants who subsequently tested qPCR positive or not. Ct value during the first 15 days of breakthrough infection in all qPCR positive participants (**A**), and in participants with an asymptomatic course of infection (**B**). Ct values in participants who were symptomatic (dark grey) or asymptomatic (light grey) at time of sampling (**C**). Ct; cyclic threshold, qPCR; qualitative polymerase chain reaction, pos; positive *; P < 0.05, ns; P > 0.05.

Five participants, all with Ct values >30 in the initial positive sample, were qPCR negative in all follow-up samples. Six participants (10%) remained asymptomatic throughout the whole course of their infection (Figure 3B).

### High post booster antibody titers are associated to protection and reduce viral load

To assess the potentially protective effect of serum antibodies against omicron infection, we investigated the change in risk of infection with every two-fold increase in binding antibody titer. Increasing titers of WT spike specific IgG and BA.1 spike specific IgG were protective against infection with relative risk estimates adjusted for age, sex, and prior infection status of 0.71 (95% CI 0.55-0.92) and 0.73 (95% CI 0.57-0.93) per two-fold increase in antibody level, Figure 4A. Relative risk of symptomatic infection estimates were similar, with RR 0.72 (95% CI 0.55 - 0.95) per every two-fold increase in WT spike-specific IgG titer.

**Figure 4.**
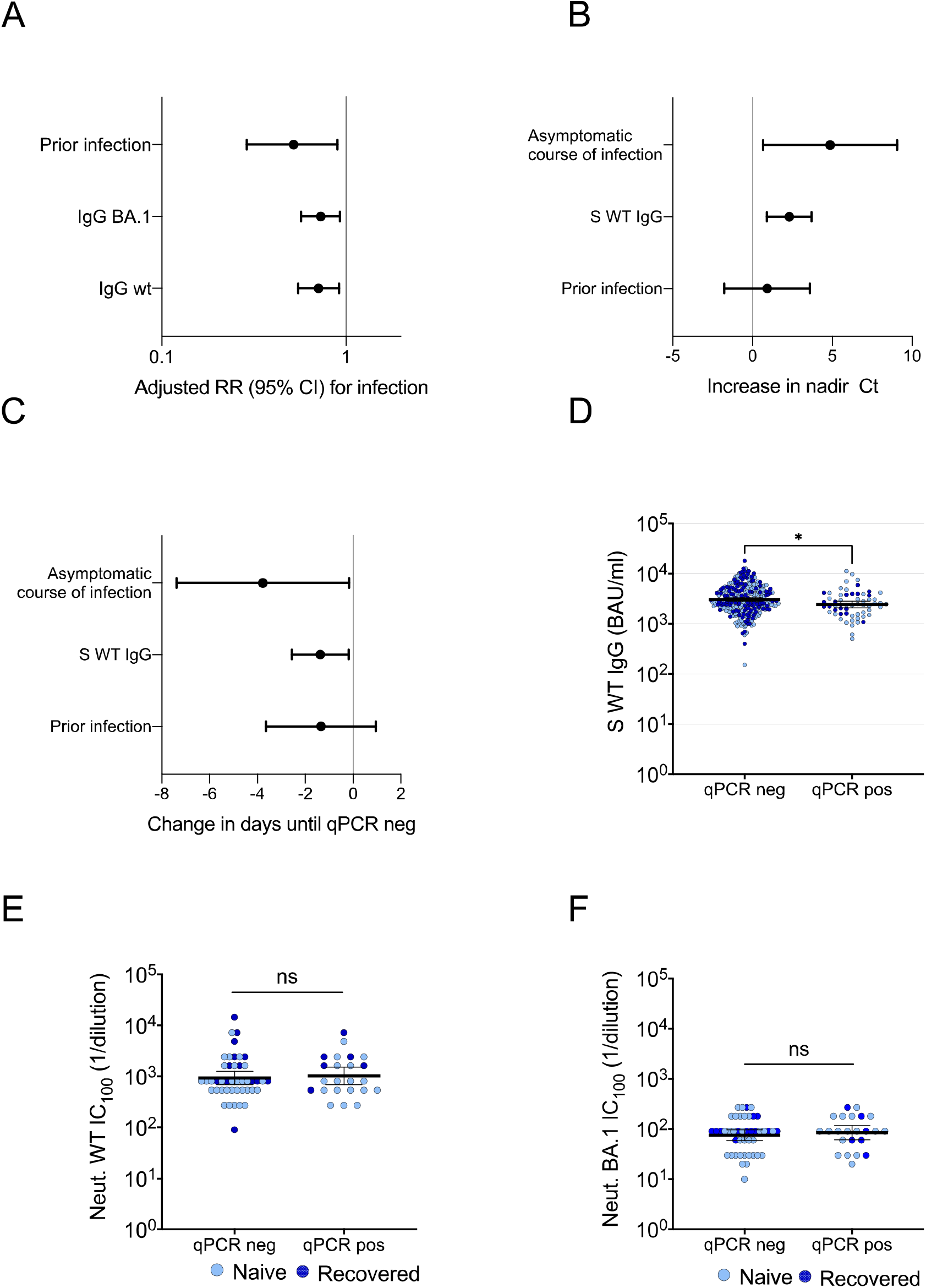
Effect of antibody levels and prior infection status on nadir Ct, time to qPCR neg and adjusted relative risks. (**A**) RR of infection in participants with prior infection vs those without and change in risk of infection per two-fold increase in post-booster antibody level, adjusted for age, sex and prior infection status/spike-specific IgG level. Estimates derived from a Poisson regression model with log offset for time in study. (**B**) Increase in nadir Ct by asymptomatic course of infection, per two-fold increase in WT spike-specific IgG levels and prior SARS-CoV-2 infection and (**C**) change in days until viral clearance (qPCR neg test) by asymptomatic course of infection, per two-fold increase in WT spike-specific IgG levels and prior SARS-CoV-2 infection. Estimates derived from a linear regression model adjusted for age, sex, asymptomatic course of infection, WT spike-specific serum IgG levels and prior infection status. (**D**) Post booster spike-specific IgG, (**E**) microneutralization titers against SARS-CoV-2 WT and (**F)** omicron BA.1 in participants that remained qPCR negative and participants that tested qPCR positive during the screening period. S; spike, WT; wild type, RR; relative risk, CI; confidence interval Ct; cyclic threshold, qPCR; qualitative polymerase chain reaction, pos; positive, neg; negative, BAU; binding antibody units; Neut; microneutralizing titer, *; p<0.05, ns; p > 0.05.

Increasing WT spike-specific IgG levels reduced peak viral load and time to viral clearance, with an increase in Ct nadir by 2.37 (95 CI 0.99 to 3.76) and a decrease in time to qPCR negativity by 1.37 (0.18 – 2.56) days per two-fold increase in WT spike-specific IgG, (fig 4B and C).

The overall difference in WT spike specific IgG levels at inclusion between participants that subsequently tested positive and those that remained negative throughout the screening period was small but significant (p=0.0012, Mann-Whitney U-test) (Fig 4D). In the subset tested with live virus neutralization, neutralizing titers against WT (figure 4E) or omicron BA.1 (figure 3F) did not differ significantly between participants with and without subsequent breakthrough infection (p>0.05, Mann-Whitney U-test).

### High viral load observed prior to onset of symptoms

More than one third, 23 of 60 (38%) participants remained asymptomatic >48 hours after first qPCR-positive sample, with a median pre-symptomatic Ct value of 28.9 (range 19.4-38).

Participants with an asymptomatic course of infection had a significantly higher nadir Ct value with a mean increase of 4.25 (95% CI 0.66-7.85), (Figure 4B), and shorter mean time to viral clearance (−3.36 days, 95% CI -6.56 to -0.15), Fig 4C. Presence of symptoms at time of sampling were correlated to a higher viral load as compared to samples from asymptomatic participants (p<0.0001, Mann-Whitney U-test) (Figure 3C). Among symptomatic participants, “common cold” symptoms dominated, fig S2A.

### Prior SARS-CoV-2 infection is associated with protection against reinfection, reduced viral load and time to viral clearance

Participants with prior SARS-CoV-2 infection had a reduced adjusted risk of testing qPCR positive (RR 0.52, 95% CI 0.29 - 0.90), (Figure 4A). Occurrence of prior SARS-CoV-2 infection was furthermore associated with a reduction in viral load, with an increase in mean nadir Ct of 2.43 (95% CI 0.13-4.73) (Figure 4B), and a clear trend towards shorter time to viral clearance by 1.35 days (95%CI -0.95 – 3.65), (Figure 4C).

### Comparisons between omicron sublineages BA.1, BA.1.1 and BA.2 breakthrough infections

Whole genome sequencing was successful in 70/71 cases with at least one sample with Ct <35, identifying 25 BA.1 (of which 19 were included in follow-up and analysis), 21 BA.1.1 (13 in follow-up and analysis), and 24 BA.2 (22 in follow-up and analysis) infections. There was a non-significant trend towards lower pre-infection anti-WT spike IgG levels among participants who subsequently became infected, and those that remained qPCR negative, regardless of omicron sublineage (Figure 5A).

**Figure 5.**
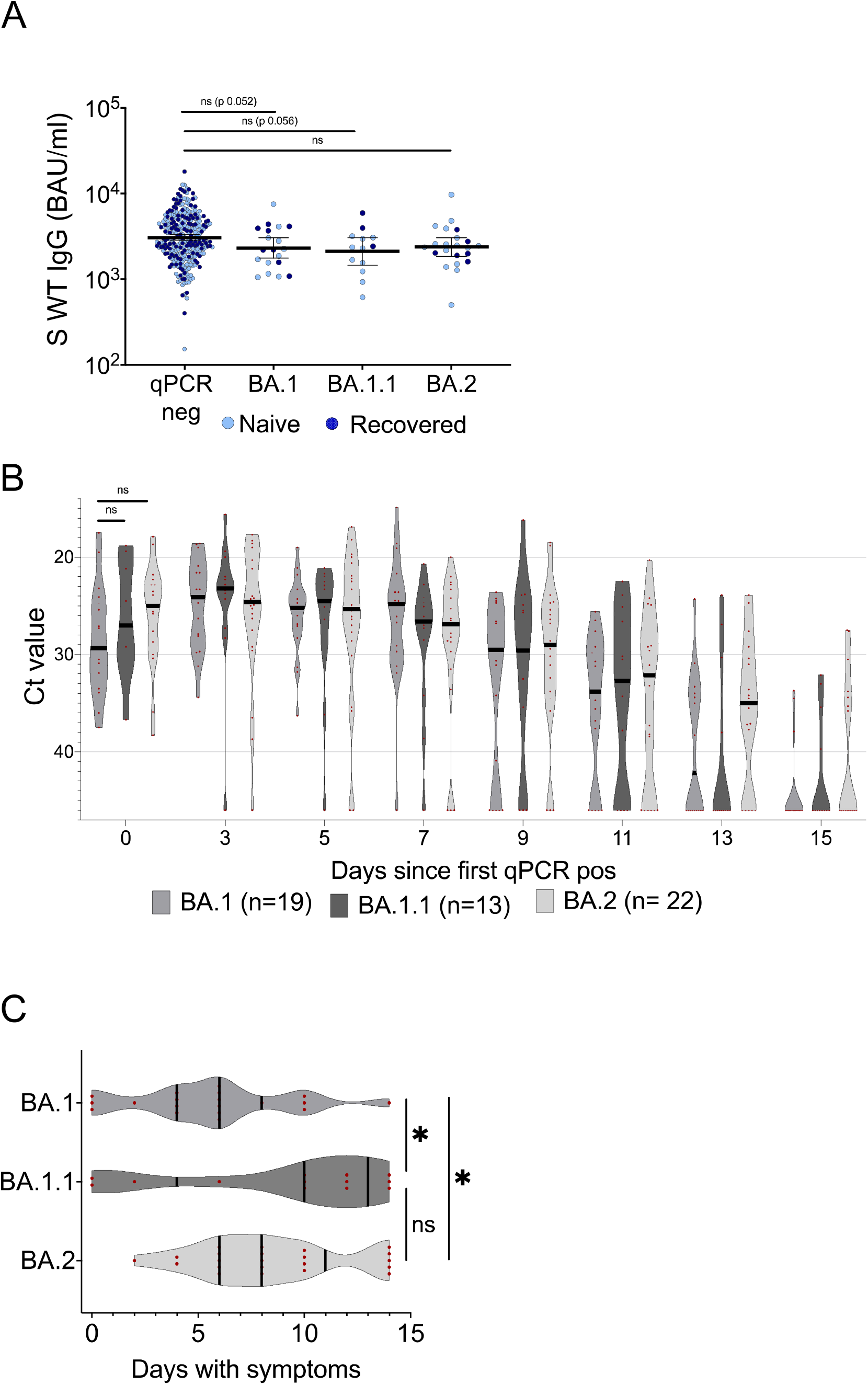
Post booster WT spike-specific IgG levels with comparisons between subsequent omicron sublineage breakthrough infections. (**A**) Post booster WT spike-specific IgG titers in participants that remained qPCR negative and in participants who tested positive with BA.1, BA1.1 or BA.2 infection during the screening period. (**B**) Ct values in qPCR positive participants the first 15 days of breakthrough infection with BA.1, BA.1.1 and BA.2. (**C**) Number of symptomatic days for participants who tested positive with BA.1, BA1.1 or BA.2 infection during the screening period. In (A), lines depict geometric mean titer and bars depict 95% confidence interval. In (B) and (C) lines depict median with IQR. Pos; positive, neg; negative, S; spike, WT; wild-type; Ct; cyclic threshold, BAU; binding antibody units, ns; p > 0.05, *; p ≤ 0.05; **p ≤ 0.01

Median Ct value of first positive sample was 29.4 in BA.1 vs. 24.5 in BA.2 infections (Figure 5B), corresponding to an approximate 100-fold higher level of viral RNA in BA.2 infected individuals early in the course of infection. These differences were however not significant (p=0.09 Mann-Whitney U-test). We could not identify any strain-dependent variations in nadir Ct or time to viral clearance (linear regression p>0.2 and p>0.6, respectively) (Figure 5B). Duration of symptoms was prolonged in BA.2- compared to BA.1- infected individuals, median duration of symptoms 8 vs 6 days (p<0.05 Mann-Whitney U-test) (Figure 5C). There were no asymptomatic cases among BA.2 infections (n=22).

Isolation of infectious viruses was successful from nine individuals (three infected with BA.1 and six with BA.1.1). Interestingly, isolation of viable viruses was possible from one sample with Ct 27 and from one sample collected at day 9, showing that samples with relatively high Ct levels and samples from late in the course of infection may contain viable virus.

### Discussion

We report a high rate of breakthrough infections (22% over a period of 4 weeks) in a cohort of HCW:s recently receiving a booster immunization and with a high rate of prior infection, supporting previous *in vitro* [25 26 28] and epidemiological [2 10] reports of omicron immune evasion. High pre-infection titres displayed a protective effect and ameliorated viral load once infected, likely contributing to the proposed short-term mRNA booster effectiveness against onward omicron viral transmission [3 10].

Stronger protection against pre-omicron SARS-CoV-2 infection with increasing antibody levels have been brought forward by several reports [5 8 29 30]. Feng et al found 80% correlation to protection against SARS-CoV-2 alpha infection at an anti-spike IgG level of 247 BAU/ml [31]. Although post booster antibody titers in our study were significantly lower in participants with subsequent omicron breakthrough infection, the difference in geometric mean titer (GMT) was small, and it is noteworthy that GMTs of both groups are 10-fold higher than the level where Feng et al found 80% protection [31] (only one participant in our study cohort had post booster anti-spike IgG titer below 247 BAU/ml). The large cumulative incidence in our material despite the high GMTs illustrates the immune evasive potential of omicron. Waning antibody titres have been reported following both primary vaccination and booster doses [32-34] and maintaining antibody levels at such high levels displayed in our material will be difficult with current vaccine strategies.

Although identification of SARS-CoV-2 RNA through qPCR is not equivalent to the detection of infectious virus, low Ct values have repeatedly been shown to correspond to viable virus in cell cultures for both omicron [15 35] and other SARS-CoV-2 variants [36 37]. Consequently, Ct values are often used as a proxy of viral load. Fall et al. reported presence of infectious omicron virus in samples obtained up to eight days after symptom onset. The presence of infectious omicron virus was similar in non-vaccinated, vaccinated and boosted individuals, implying that vaccine has little effect on viral load once infected [35], a highly relevant difference from findings concerning previous SARS-CoV-2 variants [29 38]. In line with this, we demonstrate high viral RNA levels up to nine days after first qPCR positive sample, including after symptom resolution, in the majority of omicron breakthrough infections occurring shortly after a vaccine booster dose. Together, this suggests that recently vaccinated omicron-infected individuals may transmit the virus for a longer time period than the five days quarantine from symptom onset recommended by current guidelines [39 40]. This is of particular importance in vulnerable environments such as healthcare settings. Our data are furthermore in line with a recent report demonstrating a peak in omicron viral load two-five days after symptom onset [15], where virus isolation was positive in 19% of vaccinated but not boosted infected individuals nine days after first positive qPCR test.

Asymptomatic and pre-symptomatic transmissions play important roles in transmission dynamics. Accurate estimations of the number of asymptomatic infections are key in mathematic modelling and assumptions of population immunity. No more than 10% of cases in our study had an entirely asymptomatic course of infection, contradicting an early report suggesting a high rate of asymptomatic omicron infection [41]. Importantly, although Ct values were generally higher among asymptomatic and pre-symptomatic participants, several asymptomatic cases displayed low Ct values, emphasizing the role of asymptomatic transmission also in populations with a high vaccine uptake.

This study is limited by the observational design, a cohort that is comprised of predominantly young and healthy individuals with a female dominance. Study power was not large enough to detect differences between subgroups of infected participants, such as substrains of omicron. The lack of a control group furthermore prevents us from estimating protective serological correlates. The study is however strengthened by the comprehensive screening program with high adherence to testing, thereby limiting the risk of missing transient and asymptomatic cases. The relatively large number of qPCR positive cases during a period with high circulation of BA.1, BA.1.1 and BA.2 furthermore allowed us to perform a characterization of omicron sublineage breakthrough infections. Vaccine status and prior infection were obtained from high-quality national registries, and prior infection was also determined through regular serology in the cohort, which has been followed since the start of the COVID-19 outbreak.

In conclusion, identifying potential immune correlates of protection from infection and understanding the kinetics of SARS-CoV-2 omicron shedding in vaccinated individuals is crucial to guide infection control measures and vaccination policy. We show a high incidence of omicron infection in a recently triple vaccinated HCW cohort. These breakthrough infections were associated with high viral load, which likely contributes to the global surge in cases. The very high cumulative incidence despite a recent booster vaccine dose questions the relation between the detection of vaccine induced antibodies and omicron risk prediction.

## Data Availability

All data produced in the present study are available upon reasonable request to the authors

## Acknowledgments

This work was funded by Jonas & Christina af Jochnick foundation (CT) Lundblad family foundation (CT); Region Stockholm (CT); Knut and Alice Wallenberg foundation (CT, JK); Jonas Söderquist’s scholarship (CT); Science for Life Laboratory (PN); Erling-Persson family foundation (SoH); Center for Innovative Medicine (JK, KB); and Swedish Research Council (JK). The funders above had no role in the design and conduct of the study; collection, management, analysis, and interpretation of the data; preparation, review, or approval of the manuscript; and decision to submit the manuscript for publication.

## Supplemental figures and tables

**Table S1.** Work related SARS-CoV-2 exposure divided in type of work between study participants that tested qPCR positive and not at inclusion or in the study. Pos; positive

| Type of work | Tested qPCR pos at inclusion or in study (N=81) | Did not test qPCR pos (N=287) | Total (N=368) |
| --- | --- | --- | --- |
| COVID-19 patients, n (%) | 35 (43%) | 138 (48%) | 173 |
| Patient work, not designated COVID-19 wards, n (%) | 33 (41%) | 105 (36%) | 138 |
| No patient related work, n (%) | 10 (12%) | 39 (13%) | 49 |
| Data missing n (%) | 3 (4%) | 6 (2%) | 9 |

**Figure S1.**
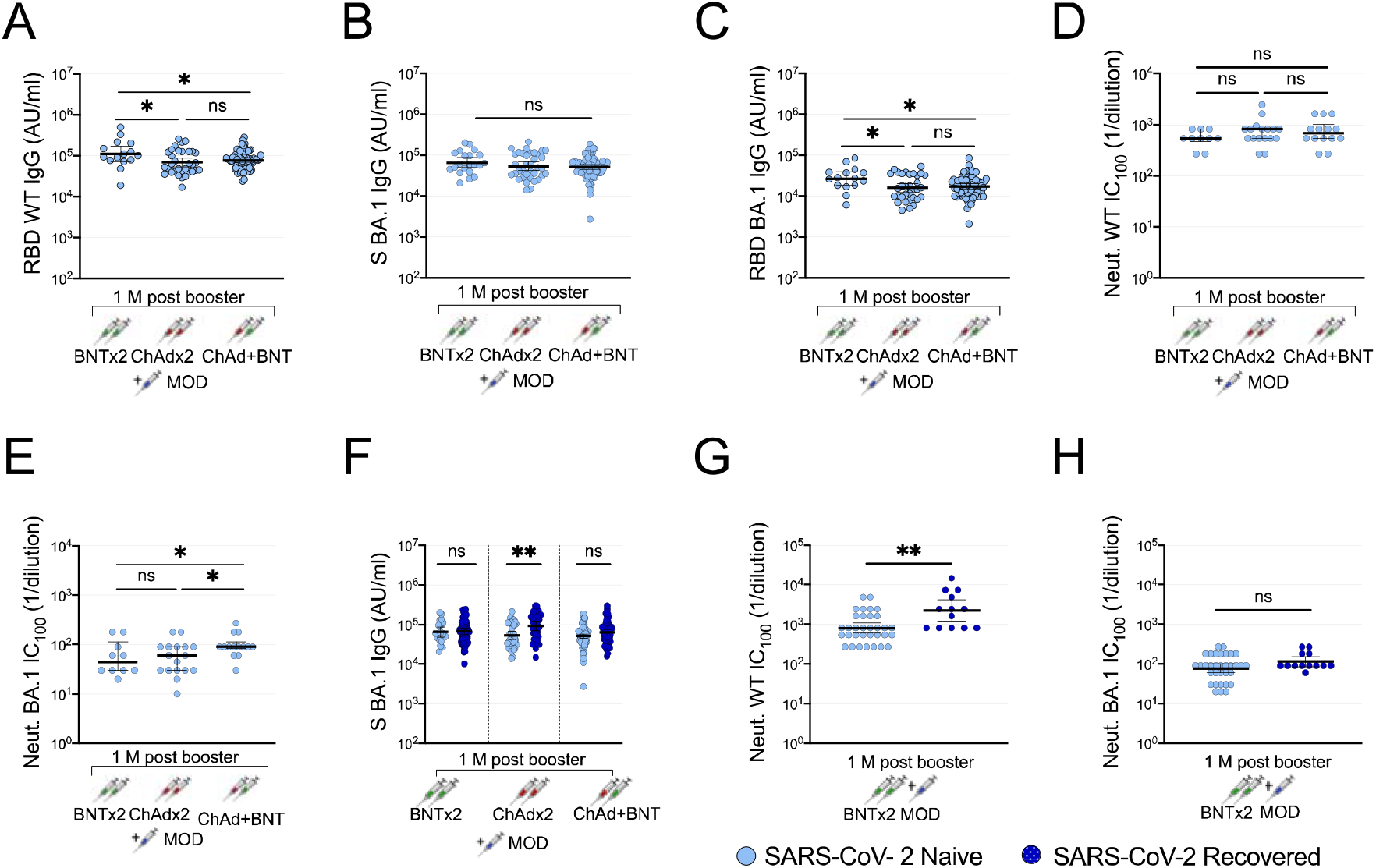
Impact of primary vaccine regimens and prior infection on booster immune responses. Anti-WT RBD IgG (**A**), cross-reactive IgG capable of binding BA.1 spike (**B**) and anti-BA.1 RBD IgG (**C**), microneutralizing titers against WT (**D**) and omicron BA.1 (**E**) one month after mRNA-1273 (MOD) booster vaccine dose in participants with primary vaccination series with two doses BNT, two doses ChAd or one dose ChAd followed by one dose BNT. Cross-reactive IgG capable of binding BA.1 Spike in SARS-CoV-2 naïve (light-blue dots) and recovered (dark-blue dots) participants one month after MOD booster vaccine dose in participants with primary vaccination series with two doses BNT, two doses ChAd or one dose ChAd followed by one dose BNT (**F**). Microneutralizing titers against WT (**G**) and omicron BA.1 (**H)** one month after MOD booster vaccine dose in participants with primary vaccination with two doses BNT. IgG titers are presented as arbitrary units (AU)/ml and microneutralizing titers are presented as lowest neutralizing dilution (1/Y). Lines depict geometric mean titers and bars depict 95% confidence interval. S; spike, WT; wild-type, Neut; microneutralizing titer, AU; arbitrary units; BNT; BNT162b2 mRNA vaccine,ChAd; ChAdox1 n-CoV 19 vaccine, MOD; mRNA-1273 vaccine, M; months, ns; P > 0.05, *; P≤0.05, **; P ≤ 0.01, ***; P ≤ 0.001, ****; P ≤ 0.0001

**Figure S2.**
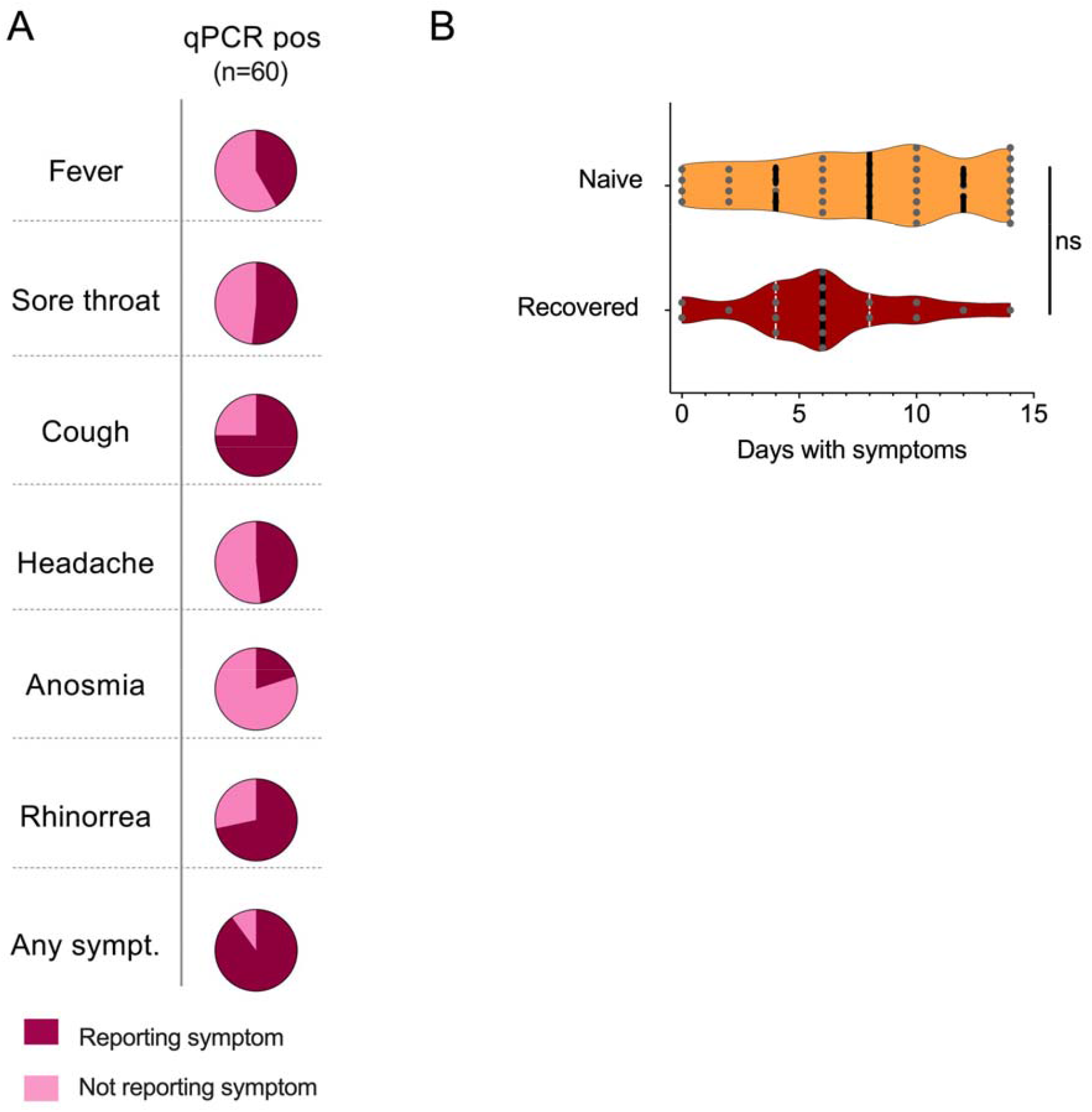
Self-reported symptomatology during omicron breakthrough infections. **(A)** Reported symptoms and prevalence of such in qPCR positive participants with (purple) and without (pink) fever, sore throat, cough, headache, anosmia and rhinorrea. (**B**) Number of symptomatic days among participants with (orange) and without (dark red) prior infection. qPCR; qualitative polymerase chain reaction, pos; positive, sympt; symptom, ns; p > 0.05.

